# Opportunistic PSA-free prostate cancer screening utilising biparametric MRI (VISIONING)

**DOI:** 10.1101/2023.09.07.23295039

**Authors:** C. Wetterauer, M. Matthias, H. Pueschel, A. Deckart, L. Bubendorf, A. Mortezavi, E. Arbelaez, D.J. Winkel, T. Heye, D.T. Boll, E. Merkle, S. Hayoz, H.H. Seifert, C.A. Rentsch

## Abstract

**Background:** Literature suggests that prostate MRI exhibits better predictive capabilities compared to prostate-specific antigen (PSA) in detecting prostate cancer (PCa). Based on this, our present study investigates biparametric MRI (bpMRI) as a PSA-independent screening tool for PCa.

**Objective:** The primary endpoint was to assess the efforts and effectiveness of identifying 20 participants with clinically significant prostate cancer (csPCa) using bpMRI.

**Design, Setting, and Participants:** Biopsy-naïve men aged over 45 years were included. All participants underwent 3 Tesla bpMRI, PSA and digital rectal examination (DRE). Targeted-only biopsy was performed in participants with a suspicious lesion (PI-RADS≥3). Men with a negative bpMRI but suspicious DRE or elevated PSA had template biopsies. Pre-intended protocol adjustments were made post-interim analysis for PI-RADS 3 lesions: follow-up mpMRI after 6 months, biopsy only if lesions persisted or upgraded.

**Outcome Measurements and Statistical Analysis:** Biopsy results underwent comparison using Fisher’s exact test and univariable logistic regression to pinpoint prognostic factors for positive biopsy.

**Results and Limitations:** A total of 229 participants were enrolled in this analysis. Among these, 77 displayed suspicious PI-RADS lesions. A total of 79 participants underwent a biopsy. PCa was detected in 29 participants, of whom 21 had csPCa.

BpMRI detected all 21 csPCa, whereas PSA and DRE missed 66.7%. Protocol adjustment led to a 54.6 % biopsy reduction in PI-RADS 3 lesions. Overall, 10.9 bpMRIs were needed to identify one participant with csPCa. A major limitation of the study is the lack of a control cohort undergoing template biopsies.

**Conclusions:** Opportunistic screening utilising bpMRI as primary tool reveals participants with csPCa that traditional methods might overlook, even at low PSA levels.

**Patient summary:** Screening with bpMRI and targeted biopsy identified csPCa in every 11^th^ man, regardless of PSA levels. Preselecting patients based on PSA > 1 ng/ml and positive family history of PCa as well as other potential blood tests may further improve the effectiveness of bpMRI in this setting.

## Introduction

Prostate cancer (PCa) is the most common cancer and the second cause of cancer death in the male population in Switzerland. The decline in PCa-related deaths in recent years has been attributed to opportunistic PCa screening based on prostate-specific antigen (PSA) [1].

Without there being any recommended cancer screening at present, the Council of the European Union has recently proposed to consider the feasibility and effectiveness of implementing organised prostate cancer screening with appropriate management on the basis of PSA and magnetic resonance imaging (MRI)[2].

Large screening programs, such as the European Randomized Study of Screening for Prostate Cancer (ERSPC) and the US Prostate, Lung, Colorectal, and Ovarian Cancer Screening Trial (PLCO), have shown a reduction of up to 30% in PCa-specific mortality [3]. The ERSPC found that 570 men had to be screened, and 18 patients had to be diagnosed to prevent one PCa-related death [4].

The ERSPC and PLCO trials have established a PSA range of 3-4 ng/ml as an indication for prostate biopsy, although using PSA as a single screening parameter has been found to be suboptimal. This is supported by reports demonstrating an area under the curve of 0.52 – 0.631 using a receiver operating characteristic (ROC) curve for PSA as a screening tool for PCa [5], [6], [7]. Furthermore, the STHLM3 model (≥10% risk) has detected PCa ISUP ≥ 2 in 9% and 23% of men with PSA ranges of 1-2 ng/ml and 2-3 ng/ml, respectively [8]

The literature suggests that prostate MRI exhibits superior performance compared to PSA for PCa detection with areas under the curve of 0.52-0.631 using ROC. The use of multiparametric MRI (mpMRI) or biparametric MRI (bpMRI) reduces negative biopsies and leads to the detection of significantly more csPCa [9], [10]. Moreover, the negative predictive value (NPV) for PCa detection with MRI ranges between 87.9% and 97.5% [11]–[15]. Despite higher initial costs compared to PSA measurement alone, screening with MRI has therefore the potential to improve PCa screening.

In this study, prospectively investigating the performance of a purely bpMRI-based opportunistic PCa screening programme, our primary outcome was the measurement of its effectiveness, represented in defined parameters. Moreover, we aimed to identify necessary adaptations to improve it.

## 1. Participants and methods

### 1.1 Ethics

The study has been conducted in compliance with the Swiss Association of Research Ethics Committees (EKNZ Nr. 2018-01965) and has been registered at clinicaltrials.gov under the number NCT03749993.

### 1.2 Study design

This study was designed as a single-centre, prospective cohort study, recruiting participants from between 01/2019 and 05/2023. The study population consisted of men over 50 years of age (over 45 years for individuals of African ancestry or with a family history of prostate cancer) who had not undergone a previous prostate biopsy and had an estimated life expectancy of more than 10 years. Participants with an acute urinary tract infection (defined as voiding symptoms and pathologic urinalysis upon clinical visit), with an NIH-Chronic prostatitis symptom index (NIH-CPSI) score of ≥ 19 or an international prostate symptom score (IPSS) ≥ 20 were treated and followed up urologically but were excluded from study participation. In addition, participants with contraindications to MRI were also excluded. All participants underwent biparametric MRI (bpMRI) of the prostate using a single 3T scanner (MAGNETOM Prisma, Siemens Healthineers, Erlangen, Germany) at the Department of Radiology of the University Hospital Basel. The bpMRI protocol used is detailed in Supplementary Table 1. T2-weighted turbo-spin echo (TSE) and diffusion-weighted imaging (DWI) series were used for lesion assessment, with reporting radiologists adhering strictly to the PI-RADS (Prostate Imaging-Reporting and Data System) v2.1 guidelines to ensure equal reporting standards for each report [16].

### 1.3 Interim analysis

An interim analysis to evaluate the performance of the study protocol was planned after a first screening round (Phase I) with the intention of being able to make amendments to improve the protocol. Interim analysis was performed after the detection of clinically significant prostate cancer in five participants. The effectiveness of the study protocol was assessed by measuring the detecting rates of PCa for bpMRI in comparison to PSA and DRE.

### 1.4 Indications for biopsy and protocol amendments

In Phase I, PI-RADS 3 lesions or higher were considered suspicious and were followed by targeted-only biopsy upon detection. MRI reports classified as PI-RADS 1 and 2 were considered clinically irrelevant and were not biopsied unless the participant had an abnormal DRE and/or an elevated PSA ≥ 10.0 ng/ml. In that case, a template biopsy was performed.

As a result of the interim analysis, protocol amendments for Phase II included a re-assessment of initial PI-RADS 3 lesions in bpMRI through a follow-up mpMRI (multiparametric MRI) after 6 months. In case of persistent or upgraded of lesions, targeted prostate biopsy was performed. Moreover, in addition to suspicious DRE and or (PSA) exceeding 10 ng/ml, PSA density ≥ 0.15 ng/ml^2^ was implemented as an indication for template biopsy in the absence of a PI-RADS lesion ≥ 3 (see Fig. 1). The protocol amendments resulting from our interim analysis (Phase I) were reviewed and approved by the Swiss Association of Research Ethics Committees.

**Figure 1:**
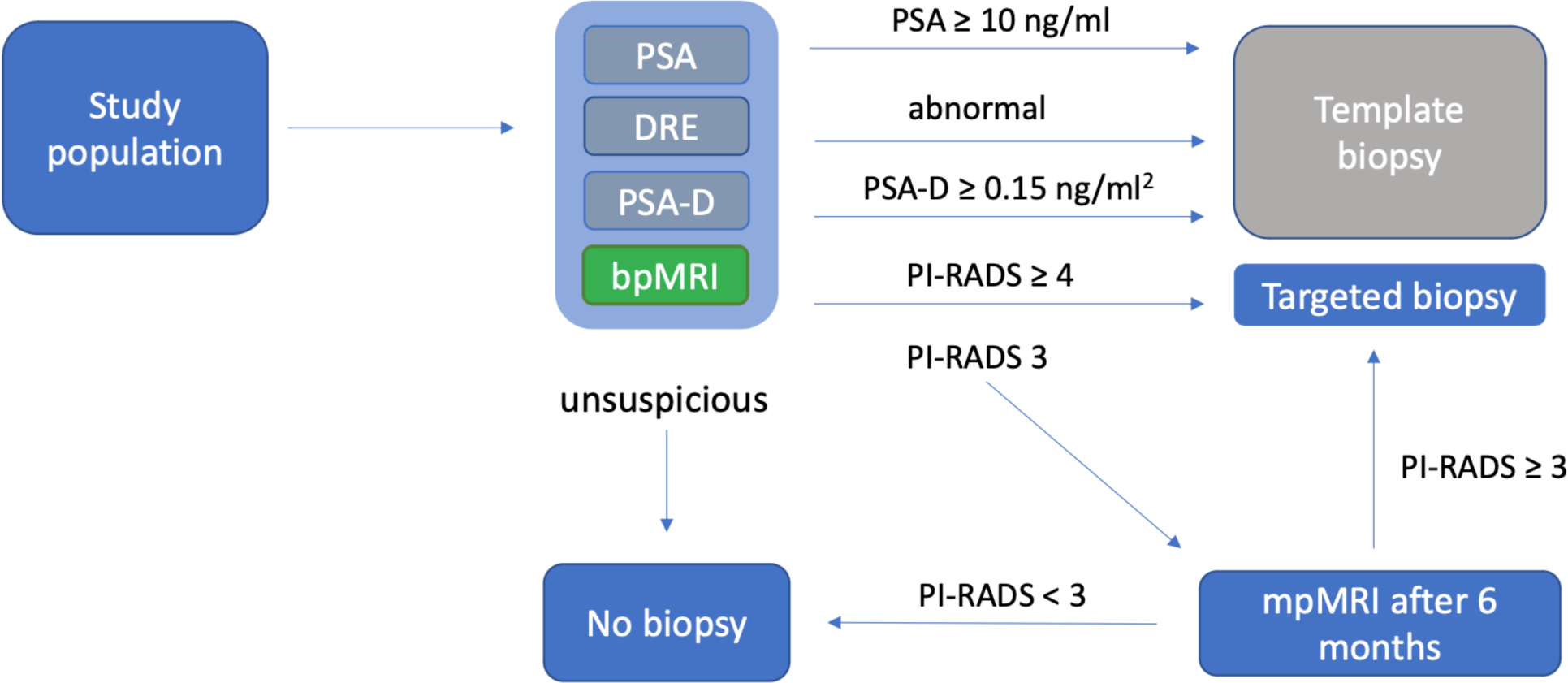
Indications for biopsy (template or targeted) and follow-up mpMRI (bpMRI = biparametric MRI; mpMRI = multiparametric MRI; PSA = prostate specific antigen; DRE = digital rectal examination; PSA-D = PSA-density)

### 1.5 Biopsy procedure

At the commencement of our study, transrectal MRI/US-fusion biopsies were performed under local anaesthesia in 36 participants. A commercially available real-time virtual sonography (RVS) system (Hitachi Medical Corporation, Tokyo, Japan) in combination with magnetic position sensors (3D Guidance Trakstar, Ascension) was used for targeted biopsies.

Subsequently, we transitioned to a transperineal approach using a robotic-assisted MRI/US-fusion biopsy system (Monalisa, Biobot Surgical Ltd, Singapore) under general anaesthesia in 39 participants. For both approaches, at least 2 cores per targeted lesion were obtained.

When indicated, a template biopsy with 12 cores equally distributed over the prostate was performed.

### 1.6 Histopathologic reporting

Gleason score and International Society of Urological Pathology Grade (ISUP) were reported for each cancer-positive biopsy. Gleason scores 3+3=6 and ISUP 1 were considered clinically non-significant PCa (cnsPCa). A Gleason score of 3+4=7 or above and ISUP 2 or above would be clinically significant PCa (csPCa).

### 1.7 Analysis and statistical methods

Categorical data were summarised using frequency and percentage, and continuous data using median and range. Biopsy results were compared between subgroups using Fisher’s exact test. Univariable logistic regression was applied to identify possible prognostic factors for positive biopsy. The significance level of 5% was used for all statistical tests with no adjustment for multiple testing, due to the exploratory nature of these analyses. All analyses were performed using SAS 9.4 (SAS Institute, Cary, NC, USA) and R 4.2.1 (The R Foundation; www.r-project.org).

## 2. Results

### 2.1 Participant enrolment and baseline characteristics

From February 2019 to September 2022, 269 participants were enrolled in our study. Finally, 229 participants were ultimately included in the analysis (Figure 2). Of the 79 participants who underwent a biopsy, all attended follow-up.

**Figure 2:**
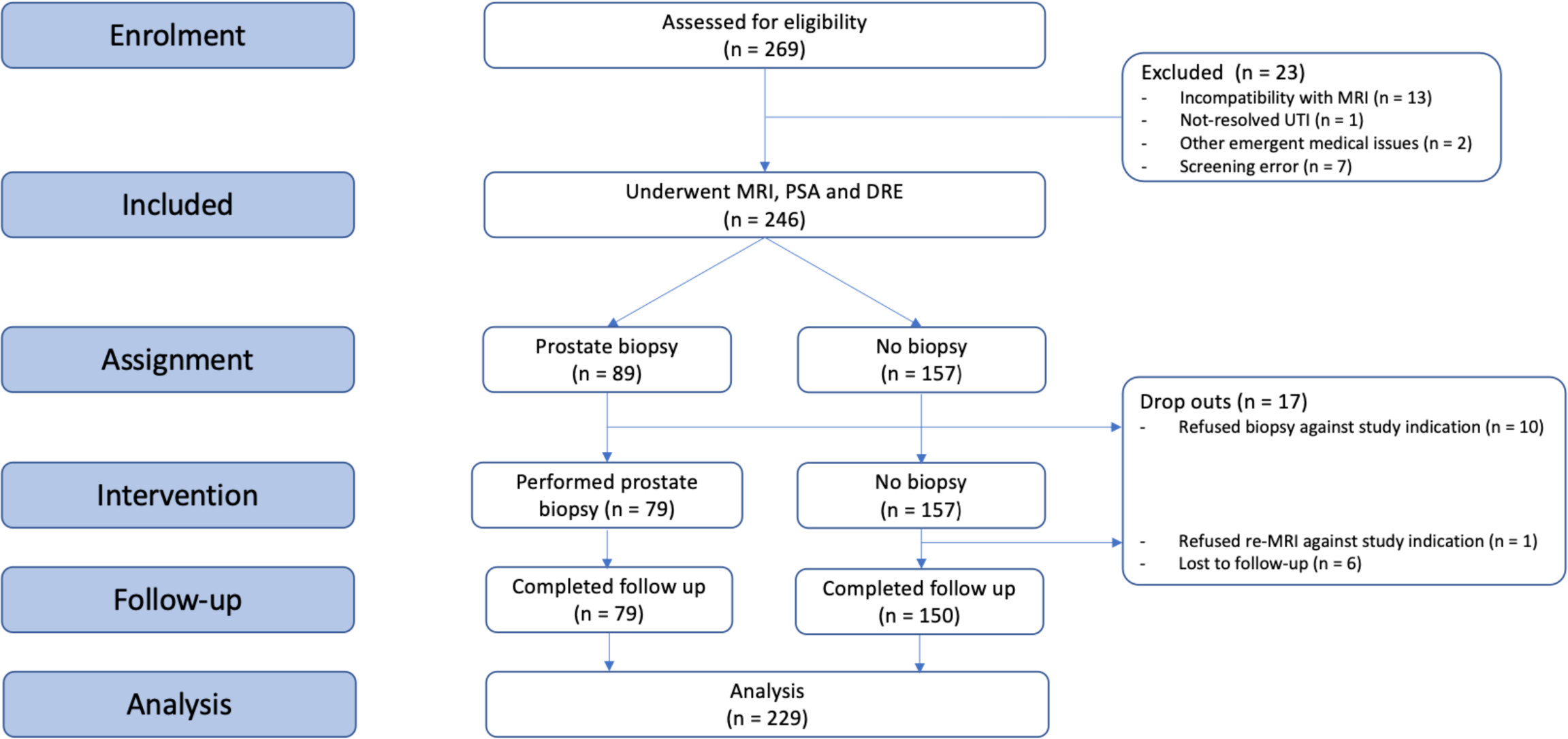
CONSORT/TREND flow diagram of the VISIONING study, follow-up and analysis (according to TREND statement) (MRI = magnetic resonance imaging; PSA = prostate-specific antigen; DRE = digital rectal examination; UTI = urinary tract infection)

The study participants had a median age of 58 years [IQR, 53–64], median PSA level of 1.26 ng/ml [IQR, 0.72–2.84] and median PSA-density of 0.05 ng/ml^2^ [IQR, 0.03–0.08]. A positive family history for PCa was found in 38 participants while 106 participants had neither a PSA > 1 ng/ml nor a positive family history for PCa (Table 1).

**Table 1:** Baseline characteristics and Clinical parameters of all included participants (PSA = prostate-specific antigen; PSA-D = PSA-density; PCa = prostate cancer).

| Baseline characteristics (n=229) | median [IQR] |
| --- | --- |
| Age (years) | 58 [53–64] |
| Prostate volume (ml) | 28 [23–39] |
| PSA (ng/ml) | 1.26 [0.72–2.84] |
| PSA-D (ng/ml<sup>2</sup>) | 0.05 [0.03–0.08] |
| Clinical parameters (n=229) | n (%) |
| PSA < 1 ng/ml & negative family history for PCa | 106 (46.2) |
| Elevated PSA ($\geq 10.0$ ng/ml) | 6 (2.6) |
| Abnormal DRE | 8 (3.5) |
| Elevated PSA-D ($\geq 0.15$ ng/ml<sup>2</sup>) | 18 (7.9) |
| Positive family history for PCa | 38 (16.6) |

### 2.2 Interim analysis (Phase I)

In the first round of opportunistic screening (n=120), we excluded 12 participants leading to 108 participants included in the analysis. Forty-five participants underwent prostate biopsy of whom 17.8 % (8/45) had a PCa ISUP ≥ 2 (Supplement figure 1). The pre-defined threshold of detecting n=5 PCa ≥ ISUP 2 in Phase I has been exceeded to n=8 because of the lead time between bpMRI, biopsy and histopathological report while the opportunistic screening had continued. Half of the ISUP ≥ 2 detected in Phase I would have been missed by PSA & DRE alone.

### 2.3 Overall results (Phase I & II)

Indication for biopsy was given in 89 of the analysed participants. Seventy-nine participants received prostate biopsy after refusal of biopsy by ten participants.

Cancer was detected in 29 of the 229 (12.7%) recruited men, of whom eight (3.5%) were ISUP 1 and 21 (9.2%) were ISUP ≥ 2. Median PSA level and PSA-density were 2.02 ng/ml [IQR, 1.5–3.21] and 0.067 ng/ml^2^ [IQR, 0.049–0.104] for the participants with ISUP 1 cancer and 3.88 ng/ml [IQR, 2.77–7.53] and 0.111 ng/ml^2^ [IQR, 0.075–0.294] for the participants with ISUP 2 or higher, respectively (Table 2).

**Table 2:** Index lesions, performed biopsy and biopsy results. *The range was specifically added in this section to display the lowest PSA values in men with prostate cancer. **In case a participant had more than one suspicious PI-RADS lesion, the highest-graded lesion would count as index lesion. ***The biopsied PI-RADS 3 lesions were from Phase I (before the interim analysis) or persistent PI-RADS 3 lesions in follow-up MRI from Phase II (PSA = prostate-specific antigen; DRE = digital rectal examination; PSA-D = PSA-density; PI-RADS = prostate imaging-reporting and data system, ISUP = International Society of Urological Pathology).

|  | n (%) | median PSA<br>(ng/ml) [IQR]<br>{Range}* | median PSA-D<br>(ng/ml<sup>2</sup>) [IQR] |
| --- | --- | --- | --- |
| **Index lesion at participant level\*\* (n=229)** |  |  |  |
| PI-RADS 1 or 2 | 152 (66.4) | 1.09 [0.67-2.2] | 0.042 [0.027-0.065] |
| PI-RADS 3*** | 13 (5.7) | 1.84 [1.15-3.96] | 0.056 [0.033-0.083] |
| PI-RADS 4 | 55 (24.0) | 2.02 [0.89-3.59] | 0.056 [0.031-0.099] |
| PI-RADS 5 | 9 (3.9) | 5.67 [2.18-20.0] | 0.151 [0.054-0.612] |
| **Biopsy intervention (n=229)** |  |  |  |
| Targeted Biopsy | 77 (33.6) | 2.06 [0.91-3.88] | 0.057 [0.031-0.111] |
| Template biopsy | 2 (0.9) | 9.4 [9.05-9.75] | 0.234 [0.196-0.272] |
| No biopsy | 150 (65.5) | 1.08 [0.67-2.17] | 0.042 [0.026-0.064] |
| **Biopsy result (n=79)** |  |  |  |
| ISUP 1 | 8 (10.1) | 2.02 [1.5–3.21]<br>{0.81-4.69} | 0.067 [0.049–0.104] |
| ISUP ≥ 2 | 21 (26.6) | 3.88 [2.77–7.53]<br>{0.8-39.7} | 0.111 [0.075–0.294] |
| No cancer | 50 (63.3) | 1.44 [0.79–3.6]<br>{0.21-12.7} | 0.051 [0.026–0.087] |

All 29 PCas were detected by bpMRI screening, while 22 would have been missed by PSA and DRE. Of these 22 cancers that classical screening would have missed, 14 (63.6%) were ISUP ≥ 2 (Figure 4).

**Figure 4:**
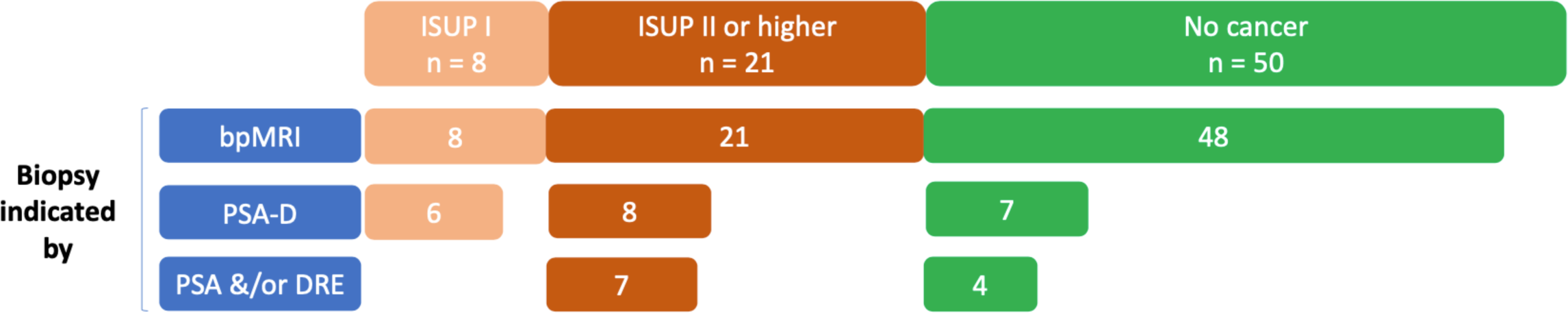
Histopathologic results (absolute numbers) overall, indicated by bpMRI, PSA-D and PSA &/or DRE. (ISUP = International Society of Urological Pathology; bpMRI = biparametric MRI; PSA = prostate-specific antigen; DRE = digital rectal examination; PSA-D = PSA-density)

As a result of the protocol amendments following the interim analysis, we could reduce the number of negative biopsies in Phase I from 73.3% (33/45) to 50% (17/34) in Phase II (Table 5). More specifically, the rate of negative biopsies indicated by MRI was reduced in Phase II from initially 72.7% (32/44) to 48.5% (16/33) by not performing an immediate biopsy in men with a PIRADS 3 score on bpMRI. The number of opportunistic screening MRIs to detect one ISUP ≥ 2 decreased from 13.6 to 9.3 while increasing the biopsy detection rate for ISUP ≥ 2 from 17.8% (8/45) to 38.2% (13/34).

We calculated detection rates for the scenario of participants with a PSA below 1 ng/ml and negative family history for PCa not being subjected to further assessment with bpMRI. Notably, by doing so, the detection of any PCa ISUP ≥ 2 would not have been compromised. We found that the number of MRIs required to detect one ISUP ≥ 2 could have decreased from 10.9 (229/21) to 5.9 (123/21). Consequently, for the entire cohort, 2.9 (61/21) targeted biopsies would have been necessary to detect one ISUP ≥ 2 (Table 4).

**Table 4:** Absolute numbers (n) of biopsies performed, biopsy results, MRIs carried out and number of MRIs needed to detect one ISUP ≥ 2 (MRI/ISUP ≥) in our cohorts of Phase I, Phase II and overall. *and/or positive family history of prostate cancer

|  | Phase I (n=108) | Phase II (n=121) | Overall (n=229) |  |
| --- | --- | --- | --- | --- |
| | Any PSA | Any PSA | Any PSA | PSA ≥ 1* |
| Biopsy performed | 45 | 34 | 79 | 61 |
| csPCa | 8 | 13 | 21 | 21 |
| cnsPCa | 4 | 4 | 8 | 6 |
| Neg. biopsy | 33 | 17 | 50 | 34 |
| MRI | 108 | 121 | 229 | 123 |
| **MRI / ISUP ≥ 2** | **13.6** | **9.3** | **10.9** | **5.9** |
| MRI / PCa | 9.1 | 7.1 | 7.9 | 4.6 |
| **Biopsy / ISUP ≥ 2** | **5.6** | **2.6** | **3.8** | **2.9** |
| Biopsy / PCa | 3.8 | 2 | 2.7 | 2.3 |

## 3. Discussion

In this study, we present the outcomes of a PSA-independent, bpMRI-based, opportunistic screening programme for PCa, comparing its effectiveness in contrast to classical screening methods.

The study encompassed 229 participants in the final analysis, and detected 21 men with ISUP ≥ 2 in the biopsy group of 79 individuals, resulting in a 26.7% detection rate for ISUP ≥ 2. The PSA-independent screening strategy using bpMRI detected all 21 men with ISUP ≥ 2, whereas classical screening would have missed 66.6% (14/21) of ISUP ≥ 2.

While adaptations were made to our study protocol after the interim analysis, the negative biopsy rate indicated by bpMRI could be reduced by 24.2% (from 72.7% down to 48.5%). However, an even more restrained strategy when indicating biopsy for PI-RADS 3 lesions does not appear to be particularly effective, as the detection rates of ISUP ≥ 2 in Phase II for PI-RADS 3 and PI-RADS 4 lesions were 25% (1/4) and 36% (9/25), respectively. As a computed result, 11 (229/21) participants had to partake in our opportunistic screening to detect one ISUP ≥ 2. The performance of opportunistic screening by bpMRI could be further improved by omitting bpMRI screening in patients with a PSA < 1 ng/ml and a negative family history for PCa. This would have led to a reduction of MRIs by 46.3% (106/229) and to a reduction of the negative biopsy rate by 7.6% (down to 55.7%) and a reduction of the diagnosis of ISUP 1 by 32% (34/50) in the whole cohort. Analysing Phase II of the study under the same assumption, only 5.9 bpMRIs and 2.9 biopsies would have been needed to detect one ISUP ≥ 2. Of note, the median PSA-density of ISUP ≥ 2 in our study was 0.111 ng/ml2, and as such, below the guidelines-suggested yet debated cut-off of 0.15 ng/ml^2^ [17].

By comparison, in the adjusted STHLM3 cohort (adjusted for age, PSA, PSA-naivety, family history and previous negative biopsy using propensity scores to match the Current Prostate Cancer Testing cohort), 53.7 participants had to be screened to detect one ISUP ≥ 2 with a biopsy rate without cancer detection of 62% [19].

While the performance of bpMRI in our setting seems very promising, the price and availability of a 3 Tesla MRI vary strongly between countries. Finally, each individual health system will have to weigh the availability and costs of MRI against more easily available and probably less expensive blood tests.

Moreover, we emphasize the financial and temporal benefits associated with bpMRI in comparison to multiparametric magnetic resonance imaging (mpMRI) when used for screening purposes with a reduced MRI scan time and omission of gadolinium-containing contrast agents. Related calculations conducted by Porter K. et al. have demonstrated that bpMRI outperforms mpMRI in terms of time and cost considerations, while still ensuring high standards of diagnostic precision [20]. Additionally, other studies have reported similar diagnostic accuracy between the two techniques [14], [21].

Regarding a pontential selection bias, as we conducted an opportunistic screening and not a screening-by-invitation programme, men with a positive family history of PCa might have been more likely to participate in this study. However, the number of participants with a positive family history of PCa was limited to 16.6%. Another evident limitation of our study protocol is the absence of a control cohort. Considering the high NPV of MRI for the detection of ISUP ≥ 2, we weighed the biopsy-related morbidity in a control cohort against the informational benefit. Therefore, we decided against the template biopsy control group. Moreover, the non-blinding of the PSA value for participants might have led to the decision to not undergo biopsy in some men with very low PSA values and PI-RADS 3 and 4 lesions.

In a recent study conducted by Moore C.M. et al. [22], a protocol similar to ours was utilized, but in a different setting. While we conducted our research under an opportunistic screening framework, they employed a screening-by-invitation approach. Remarkably, despite these differing contexts, our findings converged. This consistency between both studies underscores the robustness and potential generalizability of our results, irrespective of the screening method employed. Their conclusions, parallel to our own, highlight the significance of MRI in screening for prostate cancer, further strengthening the validity and relevance of our research. In order to answer the question whether the good performance of bpMRI also translates into better survival of men with csPCa, larger and maybe population-based screenings are required. While csPCa so far equals ISUP ≥ 2, new definitions for csPCa might be required as ISUP grade group II PCa patients with a low PSA may still profit from active surveillance instead of undergoing definitive radiotherapy or surgery.

## 4. Conclusions

This pilot study applying non-PSA triggered biparametric MRI as a sole opportunistic screening tool for prostate cancer demonstrates superior and promising effectiveness for bpMRI followed by a targeted biopsy to detect ISUP ≥ 2 compared to classical screening methods. Encompassing 229 participants overall, approximately 11 and 4 men had to undergo bpMRIs and targeted biopsies, respectively in order to detect one ISUP ≥ 2. The screening protocol may profit from adjustments leading to a further reduction of the diagnosis of ISUP 1 cancers and negative biopsies. In a first step, omitting screening in men with a PSA below 1 ng/ml and negative family history of prostate cancer would reduce the number of bpMRIs and biopsies needed to detect one ISUP ≥ 2 to approximately 6 and 3, respectively.

## 5. Take home message

BpMRI of the prostate is a promising and highly sensitive tool for PCa screening and improves the detection of ISUP ≥ 2 by 200% as compared to the current screening tools PSA and DRE.

## Data Availability

All data produced in the present study are available upon reasonable request to the authors

## Author contributions

*Data access and responsibility:* Marc Matthias had full access to all the data in the study and took responsibility for the integrity of the data and the accuracy of the data analysis.

*Study concept and design:* Rentsch, Wetterauer, Matthias

*Acquisition of data:* Matthias

*Analysis and interpretation of data:* Matthias, Rentsch Wetterauer, Hayoz

*Drafting of the manuscript:* Matthias, Wetterauer, Rentsch

*Critical revision of the manuscript for important intellectual content:* Matthias, Wetterauer, Rentsch, Seifert, Pueschel, Bubendorf, Winkel, Boll, Merkle, Heye, Hayoz, Deckart, Arbealez, Mortezavi

*Statistical analysis:* Matthias, Hayoz

*Obtaining funding:* Rentsch, Seifert, Boll, Merkle *Administrative, technical or material support:* Pueschel

*Supervision:* Rentsch, Wetterauer, Seifert

*Other:* none

## Previous presentation or release information

The results of the interim analysis of this study were presented in the form of an unmoderated poster at the annual conference of the swiss association for Urology (SGU) held from the 29^th^ of September to the 1^st^ of October 2021 in Interlaken, Switzerland, as well as in the form of an abstract at the EAU 2021. Furthermore, the presentation was held during the virtual EAU 2021 conference by Marc Matthias on the 11th of July 2021.

The current results of the primary endpoint analysis were presented as an abstract and a live presentation held by Marc Matthias at the EAU 2023 in Milano, Italy as well as at the AUA 2023 in Chicago, USA.

