## Supplements for "Opportunistic PSA-free prostate cancer screening utilising biparametric MRI (VISIONING)"

### VISIONING Supplements

| | Localizer | T2w tra (TSE) | DWI | T1w tra (TSE) |
|---|---|---|---|---|
| **TA (min)** | 0.13 | 3:37 | 2:43 | 2:45 |
| **TR (ms)** | 3.51 | 7000 | 3500 | 700 |
| **TE (ms)** | 1.53 | 104 | 66 | 12 |
| **ST (mm)** | 6 | 3 | 3 | 2 |
| **Voxel size (mm)** | 1.6 x 1.6 x 6.0 | 0.3 x 0.3 x 3.0 | 0.9 x 0.9 x 3.0 | 0.4 x 0.4 x 3.0 |
| **AF** | / | 3 | / | 3 |
| **b-values (in sec/mm$^2$)** | / | / | 0,800 | / |

*Supplement Table 1: Biparametric examination protocol for the prostate. Note that T2-weighted coronal and sagittal orientations were resliced from the transversal orientation. (TA = acquisition time, TR = repetition time, TE = echo time, ST = slice thickness, AF = acceleration factor, TSE = turbo-spin echo, DWI = diffusion-weighted imaging)*

| *Overall* | | | |
|---|---|---|---|
| | **n** | **median PSA (ng/ml) [IQR]** | **median PSA-D (ng/ml$^2$) [IQR]** |
| **bpMRI positive** | 77 | 2.06 [0.91–3.88] | 0.057 [0.031–0.111] |
| PSA &/or DRE **positive** | 11 | 10.1 [5.59–16.4] | 0.353 [0.145–0.502] |
| ***Sorted by Index lesion*** | | | |
| | **n** | **median PSA (ng/ml) [IQR]** | **median PSA-D (ng/ml$^2$) [IQR]** |
| **PI-RADS 3** | **13** | **1.84 [1.15-3.96]** | **0.056 [0.033-0.083]** |
| no cancer | 8 | 1.68 [1.11-3.85] | 0.042 [0.029-0.073] |
| ISUP 1 | 2 | 2.85 [2.27-3.42] | 0.112 [0.078-0.147] |
| ISUP $\geq$ 2 | 3 | 1.98 [1.39-6.09] | 0.076 [0.061-0.234] |
| **PI-RADS 4** | **55** | **2.02 [0.89-3.59]** | **0.056 [0.031-0.099]** |
| no cancer | 36 | 1.29 [0.72-3.21] | 0.051 [0.025-0.089] |
| ISUP 1 | 6 | 2.02 [1.15-2.76] | 0.067 [0.052-0.078] |
| ISUP $\geq$ 2 | 13 | 3.06 [2.77-5.52] | 0.083 [0.056-0.135] |
| **PI-RADS 5** | **9** | **5.67 [2.18-20.0]** | **0.151 [0.054-0.612]** |
| no cancer | 4 | 1.55 [0.9-2.58] | 0.04 [0.024-0.079] |
| ISUP 1 | 0 | - | - |
| ISUP $\geq$ 2 | 5 | 20.0 [7.96-38.0] | 0.612 [0.372-0.633] |
| ***Sorted by ISUP grade*** | | | |
| | **n** | **median PSA (ng/ml) [IQR] *{Range}** | **median PSA-D (ng/ml$^2$) [IQR]** |
| **ISUP 1** | 8 | 2.02 [1.5–3.21] {0.81-4.69} | 0.067 [0.049–0.104] |
| **ISUP $\geq$ 2** | 21 | 3.88 [2.77–7.53] {0.8-39.7} | 0.111 [0.075–0.294] |

*Supplement Table 2: Median values for PSA and PSA-D sorted by subgroups.*

**The range was specifically added in this section to display the lowest PSA values in men with PCa*

*(PSA = prostate-specific antigen; PSA-D = PSA-density; bpMRI = biparametric MRI; DRE = digital rectal examination; PI-RADS = prostate imaging-reporting and data system)*

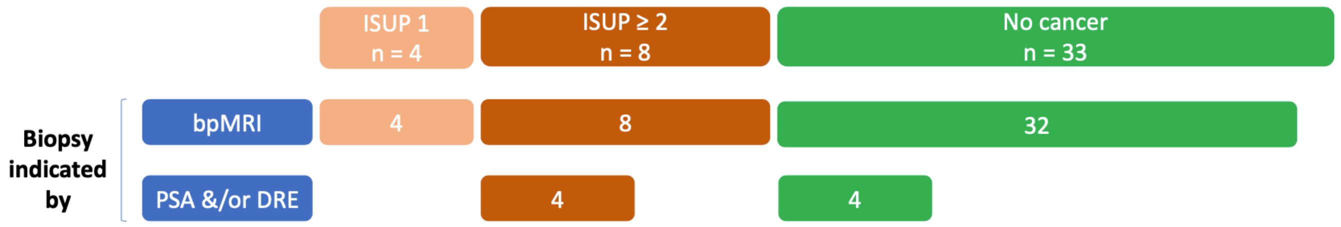


*Supplement Figure 1: Histopathologic results (absolute numbers) in the biopsy group of Phase I, indicated by bpMRI and PSA &/or DRE. Of note, the PSA-density was not part of the possible biopsy indications in Phase I. (ISUP = International Society for Urological Pathology; bpMRI = biparametric MRI; PSA = prostate-specific antigen; DRE = digital rectal examination)*

| **Subgroups of PSA and PSA-D** | | | |
|---|---|---|---|
| | ***Number of participants with assigned biopsy result (n=79)*** | | |
| ***PSA (ng/ml)*** | **No cancer** | **ISUP 1** | **ISUP ≥ 2** |
| < 3.0 | 35 | 6 | 8 |
| 3.0 – 10.0 | 13 | 2 | 9 |
| ≥ 10.0 | 2 | 0 | 4 |
| ***PSA-D (ng/ml²)*** | **No cancer** | **ISUP 1** | **ISUP ≥ 2** |
| < 0.15 | 43 | 6 | 13 |
| ≥ 0.15 | 7 | 2 | 8 |

*Supplement Table 3: Biopsy results in dependence of subgroups of PSA and PSA-Density. (PSA = prostate-specific antigen; PSA-D = PSA-density; ISUP = International Society of Urological Pathology)*

### Statistical Analysis

#### Association of PIRADS and biopsy results

| *Variable* | *PIRADS 4-5 (N=64)* n (%) | *PIRADS <4 (N=15)* n (%) | *Fisher's Exact Test* p-value (2-tailed) |
|---|---|---|---|
| Biopsy result (pos/neg) | | | 1 |
| . Negative biopsy | 40 (62.5%) | 10 (66.7%) | . |
| . Positive biopsy | 24 (37.5%) | 5 (33.3%) | . |

#### Association of PSA and biopsy results

| *Variable* | *PSA < 1 (N=20)* n (%) | *PSA >= 1 (N=59)* n (%) | *Fisher's Exact Test* p-value (2-tailed) |
|---|---|---|---|
| Biopsy result (pos/neg) | | | 0.03 |
| . Negative biopsy | 17 (85.0%) | 33 (55.9%) | . |
| . Positive biopsy | 3 (15.0%) | 26 (44.1%) | . |
| Biopsy result (categorized) | | | 0.022 |
| . Negative biopsy | 17 (85.0%) | 33 (55.9%) | . |
| . cnsPCa | 2 (10.0%) | 6 (10.2%) | . |
| . csPCa | 1 (5.0%) | 20 (33.9%) | . |

Separate by Phase:

Phase I

| *Variable* | *PSA < 1 (N=13)* *n (%)* | *PSA >= 1 (N=32)* *n (%)* | *Fisher's Exact Test* *p-value (2-tailed)* |
|---|---|---|---|
| Biopsy result (pos/neg) | | | 0.134 |
| . Negative biopsy | 12 (92.3%) | 21 (65.6%) | . |
| . Positive biopsy | 1 (7.7%) | 11 (34.4%) | . |
| Biopsy result (categorized) | | | 0.121 |
| . Negative biopsy | 12 (92.3%) | 21 (65.6%) | . |
| . cnsPCa | 1 (7.7%) | 3 (9.4%) | . |
| . csPCa | 0 (0.0%) | 8 (25.0%) | . |

Phase II

| *Variable* | *PSA < 1 (N=7)* *n (%)* | *PSA >= 1 (N=27)* *n (%)* | *Fisher's Exact Test* *p-value (2-tailed)* |
|---|---|---|---|
| Biopsy result (pos/neg) | | | 0.398 |
| . Negative biopsy | 5 (71.4%) | 12 (44.4%) | . |
| . Positive biopsy | 2 (28.6%) | 15 (55.6%) | . |
| Biopsy result (categorized) | | | 0.338 |
| . Negative biopsy | 5 (71.4%) | 12 (44.4%) | . |
| . cnsPCa | 1 (14.3%) | 3 (11.1%) | . |
| . csPCa | 1 (14.3%) | 12 (44.4%) | . |

**By PSA >1 or family history for prostate cancer:**

| Variable | PSA < 1 and no family hi (N=16) n (%) | PSA >= 1 or family histo (N=63) n (%) | Fisher's Exact Test p-value (2-tailed) |
|---|---|---|---|
| Biopsy result (pos/neg) | | | 0.004 |
| . Negative biopsy | 15 (93.8%) | 35 (55.6%) | . |
| . Positive biopsy | 1 (6.3%) | 28 (44.4%) | . |
| Biopsy result (categorized) | | | 0.006 |
| . Negative biopsy | 15 (93.8%) | 35 (55.6%) | . |
| . cnsPCa | 1 (6.3%) | 7 (11.1%) | . |
| . csPCa | 0 (0.0%) | 21 (33.3%) | . |

**Univariable logistic regression for positive biopsy**

Overall

| | Odds Ratio (95% CI) | p-value |
|---|---|---|
| PSA | 1.17 (1.01 - 1.36) | 0.040 |
| Family history of prostate cancer (yes vs no) | 2.00 (0.66 - 6.08) | 0.221 |
| Stage (II vs. I) | 2.75 (1.07 - 7.06) | 0.035 |
| PSA (categorized) (PSA ≥ 1 vs. PSA < 1) | 4.46 (1.18 - 16.89) | 0.028 |
| PI-RADS (categorized v2) (PI-RADS 4-5 vs. PI-RADS <4) | 1.20 (0.37 - 3.93) | 0.763 |

Phase I

| | Odds Ratio (95% CI) | p-value |
|---|---|---|
| PSA | 1.17 (0.95 - 1.45) | 0.134 |
| Family history of prostate cancer (yes vs no) | 1.45 (0.23 - 9.16) | 0.693 |
| PSA (categorized) (PSA ≥ 1 vs. PSA < 1) | 6.29 (0.72 - 54.86) | 0.096 |
| PI-RADS (categorized v2) (PI-RADS 4-5 vs. PI-RADS <4) | 0.81 (0.17 - 3.81) | 0.787 |

Phase II

| | Odds Ratio (95% CI) | p-value |
|---|---|---|
| PSA | 1.16 (0.93 - 1.47) | 0.194 |
| Family history of prostate cancer (yes vs no) | 1.77 (0.40 - 7.93) | 0.454 |
| PSA (categorized) (PSA ≥ 1 vs. PSA < 1) | 3.12 (0.51 - 19.04) | 0.216 |
| PI-RADS (categorized v2) (PI-RADS 4-5 vs. PI-RADS <4) | 1.61 (0.23 - 11.09) | 0.630 |
